# Public attitudes towards COVID-19 vaccines in Africa: a systematic review

**DOI:** 10.1101/2022.04.19.22274053

**Authors:** Patrice Ngangue, Arzouma Hermann Pilabré, Abibata Barro, Yacouba Pafadnam, Nestor Bationo, Dieudonné Soubeiga

**Author notes:** **Corresponding author Patrice NGANGUE, MD, MSc, Ph.D.**, Institut de Formation et de Recherche Interdisciplinaires en Sciences de la Santé et de l’Éducation, IFRISSE, Postal box: 09 BP 311 Ouagadougou 09, Ouagadougou, Burkina Faso. **Authors’ contributions** PN, FB bibliographic researches; PN, HP, FB, YP screening and data extraction data, quality assessment; PN, HP synthesis; PN, HP manuscript writing with input from NB and DS; all authors read and approved the final manuscript.

## Abstract

**Background:** As COVID-19 vaccine acquisition and deployment accelerates, tensions also increase. This review aims to identify and understand the significance of population attitudes toward COVID-19 vaccines in Africa.

**Methods:** A systematic review was conducted. Searches were conducted in MEDLINE, CINAHL, EMBASE, and Global Health databases. Database searches began on June 23, 2021, and the last search date was June 30, 2021. The methodological quality of the studies included in this review was assessed using the Mixed methods appraisal tool.

**Results:** A total of 609 articles were retrieved, and 23 met the eligibility criteria. All 23 included studies were cross-sectional. Three attitudes were identified: acceptance, reluctance, and refusal to be vaccinated. Acceptance of vaccination was motivated by confidence in the accuracy of the government’s response to COVID-19 and the fact that relatives had been diagnosed with or died from COVID-19. Reluctance was based on fear of vaccine quality and side effects, and insufficient clinical trials. Finally, refusal to be vaccinated was justified by reasons such as the unreliability of clinical trials and insufficient data regarding the vaccine’s adverse effects.

**Conclusion:** This review revealed common attitudes of African populations toward COVID-19 vaccines. The results indicate that research needs to focus more on identifying facilitators of COVID-19 vaccination. However, they also provide essential elements for health personnel in charge of vaccination to develop strategies to achieve satisfactory coverage rates

## Introduction

In December 2019, a cluster of patients presented with pneumonia caused by an unknown pathogen linked to the seafood wholesale market in Wuhan, China. Subsequently, a new coronavirus was identified by sequencing the whole genome of patient samples.^[1]^ It was named severe acute respiratory syndrome coronavirus 2 (SARS-CoV-2) by the Coronavirus Study Group of the International Committee on Taxonomy of Viruses,^[2]^ and the disease caused by the virus was named coronavirus disease 2019 (COVID-19) by the WHO.

After infecting and causing the death of thousands of persons in China, the virus has spread, reaching Italy and other European countries and the USA, with the number of confirmed new cases currently increasing every day.^[3]^ As a result, the WHO declared it a pandemic due to the widespread infectivity and high contagion rate.

Since then, the world has experienced much uncertainty due to changing COVID-19 evidence, new and emergent strains of the virus, and an ever-shifting landscape of travel bans and lockdowns. The global efforts to lessen the effects of the pandemic and reduce its health and socio-economic impact rely largely on preventive efforts.^[4,5]^ Thus, tremendous efforts by the scientific community and pharmaceutical industry-backed by governments’ support were directed towards developing efficacious and safe vaccines for SARS-CoV-2. ^[6]^ These efforts were manifested by the approval of several vaccines for emergency use, in addition to more than 60 vaccine candidates in clinical trials. Ensuring a solid understanding of, demand for, and promoting acceptance of current and forthcoming COVID-19 vaccines is critical to personal health, protecting the most vulnerable populations, reopening social and economic life, and potentially achieving population health and safety through immunity.^[7]^ COVID-19 vaccines have generated a renewed sense of hope for many devastated by deaths and livelihoods due to the disease.

However, tensions are also growing as the acquisition and roll-out of COVID-19 vaccines gain momentum. Emerging COVID-19 vaccine hesitancy is an additional concern.^[8,7]^ The World Health Organization has defined vaccination hesitancy as “the delay in acceptance or refusal of vaccines despite the availability of services.^[9]^ “ Refusal is the choice made by some people not to accept vaccination against COVID-19. Several recently conducted national, continental, and global surveys suggest that hesitancy and refusal of COVID-19 vaccines is an emerging problem.^[10,11]^ Indeed, a rapid systematic review of 126 surveys on COVID-19 vaccination intentions (covering a total of 31 countries), including 23 academic studies and 103 opinion polls published by October 20, 2020, found declining global vaccine (anticipated) acceptance, from greater than 70% in March to less than 50% in October.^[12]^ Against this backdrop, addressing current and future potential COVID-19 vaccine hesitancy is critical.

Previous work has identified several factors for vaccine reluctance. These factors include lack of trust in pharmaceutical companies, doubt about the quality of vaccines, negative perception of vaccine efficacy and convenience, pain associated with injection, or fear of injection.^[13,14]^

To our knowledge, no systematic review has explored how attitudes toward the COVID-19 vaccine have changed during the pandemic in Africa. Therefore, there is a need to identify and capture the meaning of African populations’ attitudes toward COVID-19 vaccines. In this study, attitude should be understood as a settled way of thinking about the COVID-19 vaccine ranging from negative to positive to hesitant. These findings may be significant in ensuring population adoption of the COVID-19 vaccine.

## Methods

To methodology of this review followed the guidelines of the Preferred Reporting Items for Systematic Review and Meta-Analysis (PRISMA). ^[15]^

### Eligibility criteria

The criteria for inclusion of articles were as follows:

– original research articles on COVID-19 vaccination intent;
– research with a quantitative, qualitative or mixed estimate;
– studies in which participants are the general population and specific population groups.

The exclusion criteria were:

– studies published in languages other than English and French;
– commentaries, summary documents, case studies, letters, discussion papers, posters, conference summaries, conference reports and briefs;

### Eligibility criteria

The criteria for inclusion of articles were as follows:

– original research articles on COVID-19 vaccination intent (Any direction taken in a person’s thoughts or behaviors regarding COVID-19 vaccination, whether or not it involves conscious decision making);
– research with a quantitative, qualitative or mixed-method;
– studies in which participants are the general population and specific population groups.

The exclusion criteria were:

– studies published in languages other than English and French;
– commentaries, summary documents, case studies, letters, discussion papers, posters, conference summaries, conference reports and briefs.

### Information sources

We searched PubMed / MEDLINE, CINAHL, EMBASE, and Global health databases. Literature search strategies were developed using a free and controlled vocabulary. The database searches began on June 23, 2021, and the last search date was June 30, 2021. Reference lists of included studies were also reviewed for inclusion and recent citations of included studies.

### Search strategy

The search strategy for this study was tailored to each database based on index terms, including medical subject headings (MeSH), truncations, and Boolean operators. In addition, a combination of terms for the concepts of COVID-19 (SARS-CoV-2, novel coronavirus, coronavirus), vaccine (COVID-19 vaccines, SARS-CoV-2 vaccine), attitudes (attitude, confidence, psychology, intention, psychological distress, Vaccination reluctance, acceptance, conspiracy beliefs, enablers) and Africa were used.

The complete search strategy, line by line of each database as follows:

### PubMed/Medline strategy

1. “Women” [MeSH] OR “Men” [MeSH] OR “Adult” [MeSH] OR “Health Personnel” [MeSH] OR “Students, Medical” [MeSH] OR “Young Adult” [MeSH]
2. “COVID-19 Vaccines” [MeSH]
3. “Attitude” [MeSH] OR “Trust” [MeSH] OR “Psychological Distress” [MeSH] OR “hesitancy” OR “Acceptance” OR “Conspiracy beliefs “ OR “intention” [MeSH Terms] OR “Patient Acceptance of Health Care”[Mesh] OR “willingness”
4. “Africa”[Mesh]

### Embase

1. ‘Women’ OR ‘Men’ OR ‘Adult’ OR ‘Health Personnel’ OR ‘Students, Medical’ OR ‘Young Adult’
2. ‘COVID-19 Vaccines’ OR ‘SARS-CoV-2’
3. ‘Attitude’ OR ‘Trust’ OR ‘Psychology’ OR ‘Psychological Distress’ OR ‘ Vaccine hesitancy ‘ OR ‘Acceptance’ OR ‘Conspiracy beliefs’ OR ‘COVID-19 phobia’ OR ‘Facilitators’

### CINAHL strategy

1. (MH “Women”) OR (MH “Men”) OR (MH “Students, Medical”) OR (MH “Health Personnel”)
2. (MH “COVID-19 Vaccines”)
3. (MH “Attitude”) OR (MH “Trust”) OR (MH “Psychological Distress”) OR (TI “Vaccine hesitancy”) OR (AB “Vaccine hesitancy”) OR (MH “Acceptance and Commitment Therapy”) (TI “Conspiracy beliefs”) OR (AB “Conspiracy beliefs”) OR (MH “intention”) OR (MH “motivation”) OR (TI “willingness”) OR (AB “willingness”) OR (MH “fear”) OR MH “refusal to participate”)

### Global health

1. “Women” OR “Men” OR “Adult” OR “Health Personnel” OR “Students, Medical” OR “Young Adult”
2. “COVID-19 Vaccines” OR “SARS-CoV-2”
3. “Attitude” OR “Trust” OR “hesitancy” OR “acceptance” OR “refusal” OR “willingness” OR “beliefs “ OR “Fear” OR “motivation” OR “intention”

All study alerts published in English or French before June 30, 2021, were included.

### Selection process

A citation management system (Zotero) was used to manage records exported from all electronic databases. Two independent reviewers performed article selection. A predefined selection form was developed to ensure the reliability of article selection by the two reviewers, and a pilot test was conducted based on the eligibility criteria. Both reviewers described the outcome measures after reviewing the studies to verify the relevance of the articles. Each reviewer provided strong justifications for excluding studies. A third reviewer resolved any disagreement between the two reviewers in a consensus meeting. The third reviewer was consulted to decide whether the study met the eligibility criteria for inclusion.

Titles and abstracts were used to screen out all studies first, followed by full text to screen out studies that did not meet the inclusion criteria. Database searches initially identified a total of 69 studies. After deduplication, 59 potentially relevant titles were included for title or abstract screening. After title and abstract screening, 27 articles were excluded. Finally, the full texts of the remaining 32 studies were reviewed to determine whether they met the inclusion criteria. Ultimately, 23 studies were selected and used for this review. The Preferred Reporting Items for Systematic Reviews and Metanalyses (PRISMA) diagram was used to report the study selection process (Fig. 1).

**Figure 1.**
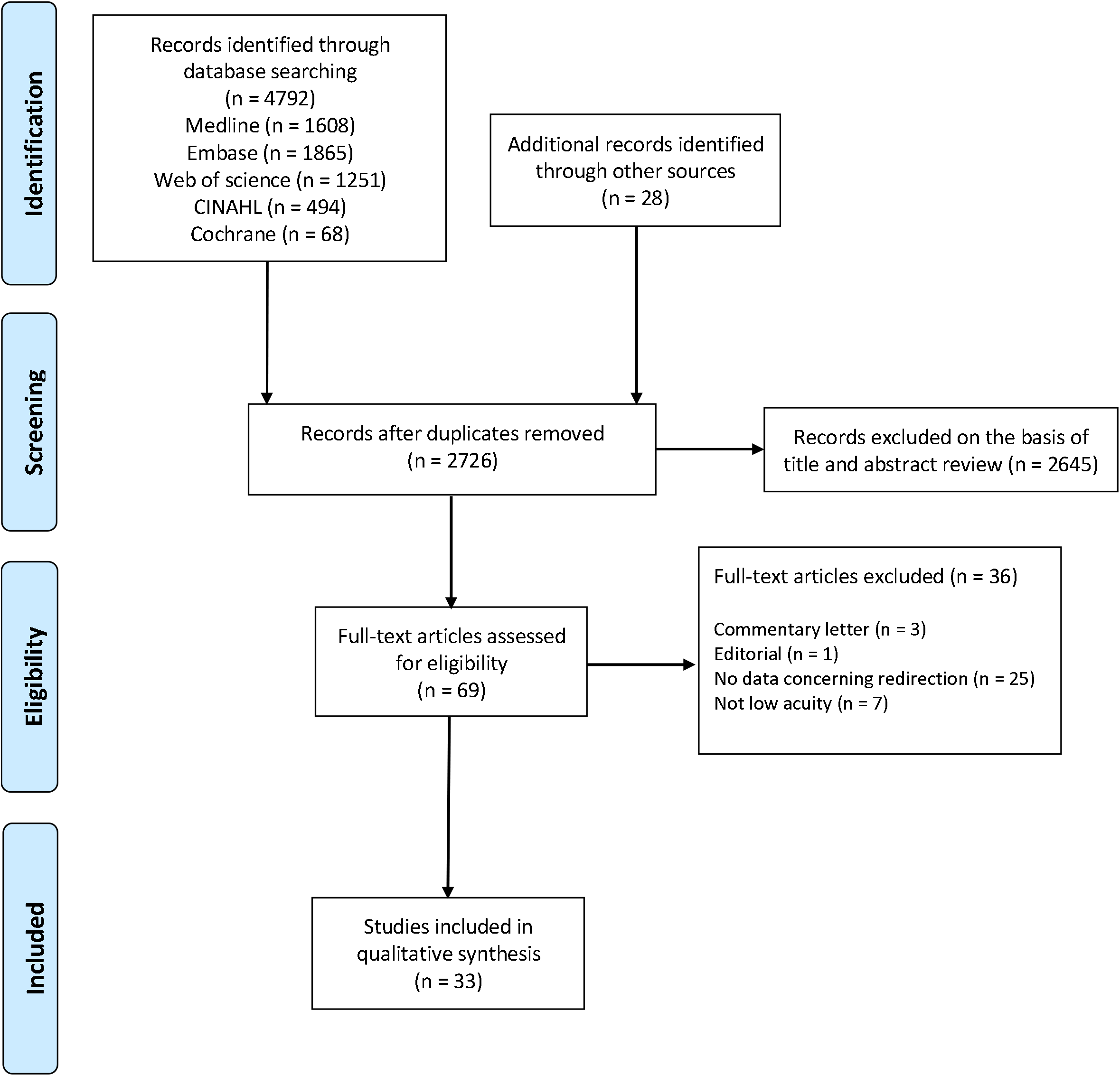
Flow diagram of included studies.

### Data collection process

The two independent reviewers completed a customized data extraction sheet for all included studies. The data extraction tables from both reviewers were matched to ensure that all key outcomes were included in the systematic review. The third reviewer was involved if conflicting information was observed during the data extraction process.

### Data item

We extracted data on the first author, study area, year of study publication, the study scope, estimate, participants, author, rate of acceptance, reasons of acceptance/intention to accept. Some of these extracted data have been presented in tabular form in the “Results” section.

### Study risk of bias assessment

The methodological quality of the studies included in this review was assessed using the MMAT (Mixed methods appraisal tool). The MMAT is a critical appraisal tool designed for mixed systematic reviews, including qualitative, quantitative, and mixed methods studies. It assesses the methodological quality of five categories of studies: qualitative research, randomized trials, quantitative descriptive studies, and mixed methods studies [16]. The MMAT Criteria List includes two triage questions and five questions per study category. In addition, the document includes indicators that explain and illustrate certain criteria. For each question, the authors answered by checking “Yes”, “I don’t know” or “No”. One author reviewed six articles, and another reviewed two articles. The authors discussed the evaluation results for all included articles with particular attention to the questions that were checked “don’t know” or “no”.

### Synthesis methods

The main findings of the studies were analyzed and summarized narratively. In addition, a systematic narrative synthesis was provided with the information presented in the text and the table to summarize and explain the characteristics and results of the included studies.

The results of the review were synthesized narratively. First, we performed a descriptive analysis of all included final studies to record their main characteristics. A narrative synthesis was then performed in which the final studies were grouped according to key attitudes.

## Results

### Studies selection

A total of 61 articles were retrieved through various searches. After removing duplicates, there were 51 articles, and 23 met the inclusion and exclusion criteria.

### Study characteristics

All 23 included studies were cross-sectional studies. The studies were carried out in Ethiopia (n = 6), Uganda (n = 3), Egypt (n = 4), Nigeria (= 2), Ghana (n = 2), Democratic Republic of the Congo (n = 1), South Africa (n=2), Tunisia (n=1) and in Libya (n = 1). Study populations were diverse including general population (n = 8), adults aged 18 to 70 (n = 1), healthcare workers (n = 9), medical students (n = 2), pregnant women attending an antenatal care clinic (n = 1), patients attending outpatient clinics (n = 1), and elementary and secondary school teachers (n = 1) (see tables 1, 2, 3).

**Table 1.**
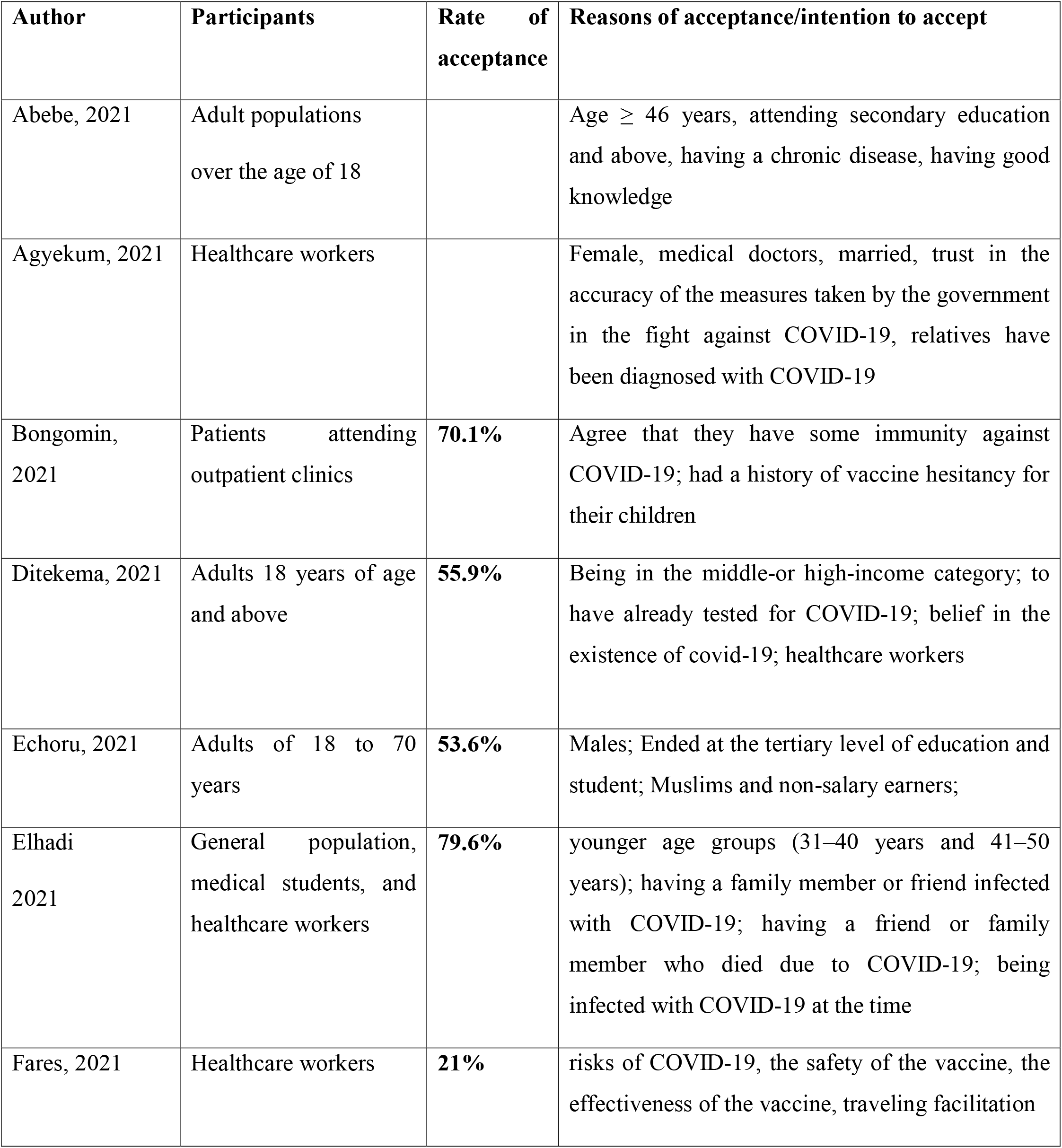
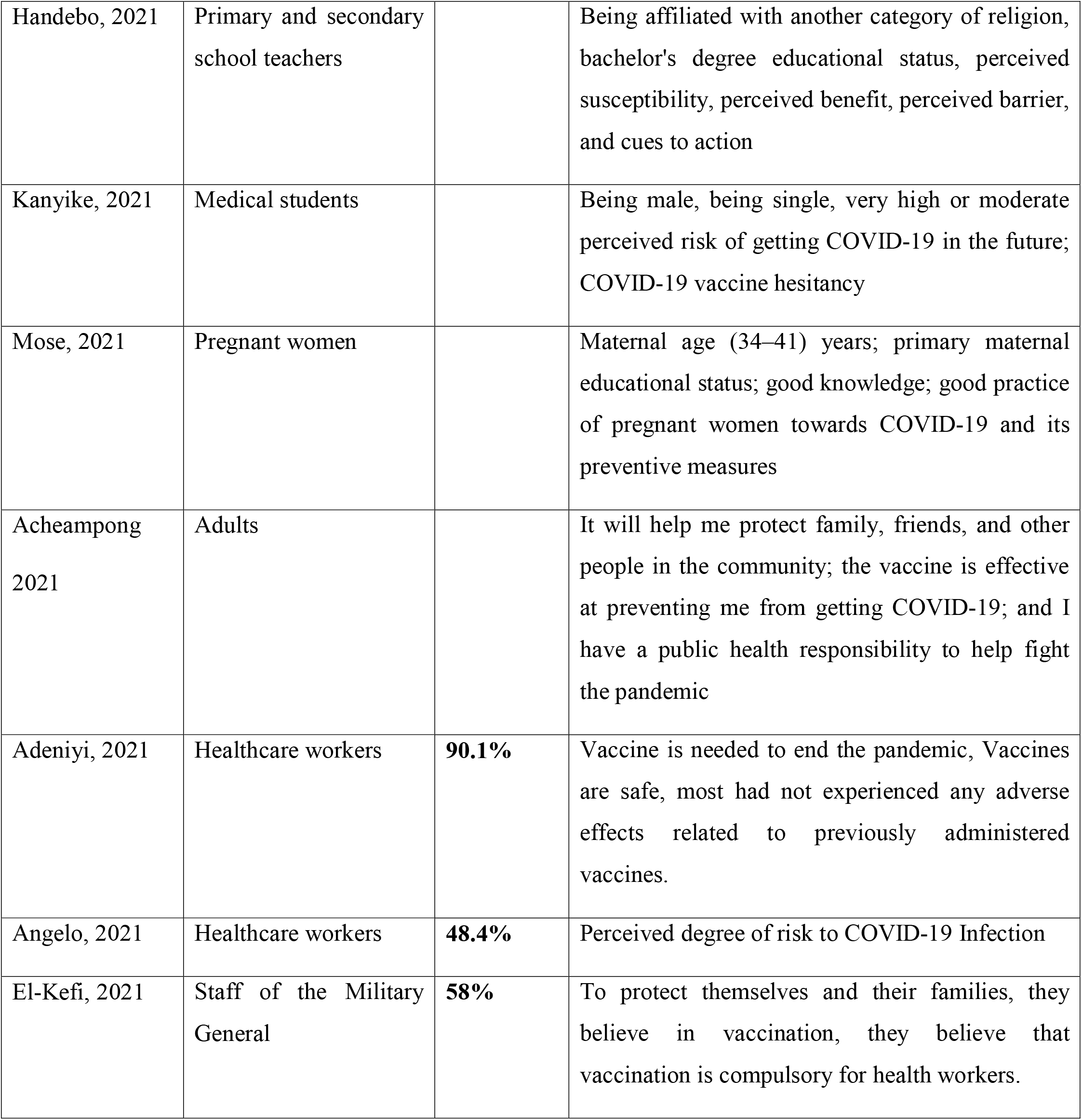
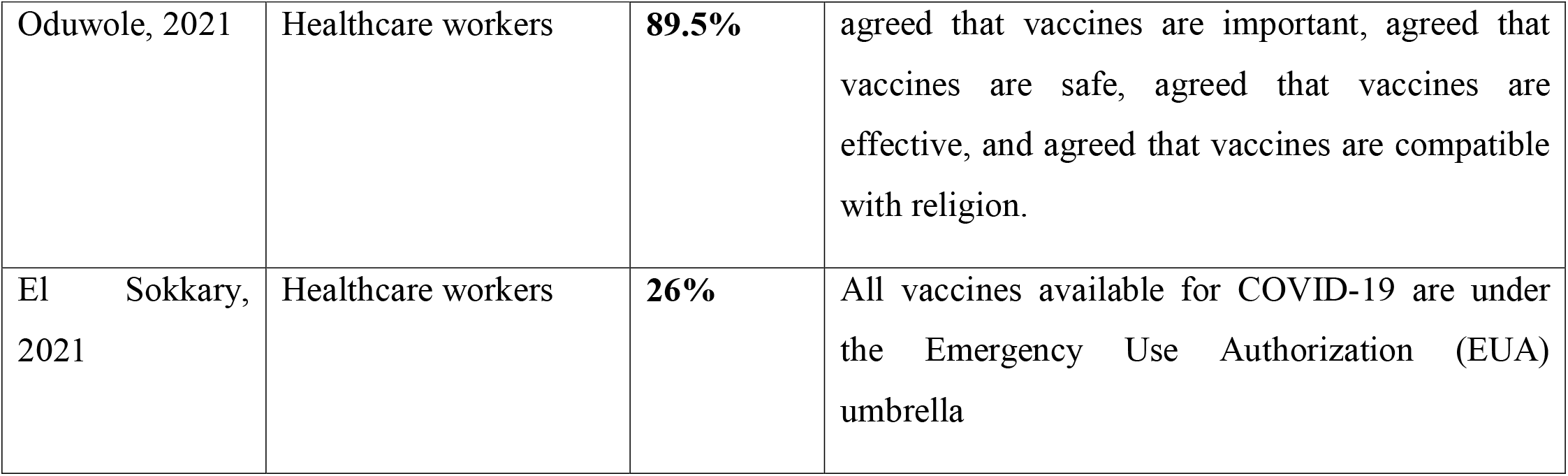
Acceptance of vaccines (rate and reasons)

**Table 2.** Hesitancy to get vaccinated (rate and reasons)

| Author | Participants | Rate of hesitancy | Reasons of hesitancy |
| --- | --- | --- | --- |
| Abebe, H, 2021 | All adult populations over the age of 18 years old | | Age ≥ 46 years, attending secondary education and above, having a chronic disease, having good knowledge. |
| Dinga, 2021 | Adults 18 years of age and above | **84.6%** | worried about the quality of the vaccine distributed or sent to Africa in general and Cameroon specifically |
| Echoru, 2021 | Adults of 18 to 70 years | **46.4%** | Aged 61–70; unemployed and pagans; unmarried group and urban dwellers |
| Fares, 2021 | Healthcare workers | **51%** | Lack of clinical trials (92.4%) and fear of the vaccine's side effects (91.4%). |
| Acheampong 2021 | Adults | **28%** | Insufficient information on the possible effects of the vaccine; uncertainty about the quality of the vaccine; and uncertainty about the effectiveness of the vaccine in preventing them from contracting COVID-19 |
| Aemro, 2021 | Healthcare workers | **45,9%** | Unclear information provided by public health authorities; low risk of contracting COVID-19 Infection; uncertainty regarding the tolerability of side effects of the vaccine |
| Amuzie, 2021 | Healthcare workers |  | younger age, marital status (single), lower income, and profession (doctor, nurse, other allied professionals), |
| Omar, 2021 | Age 18 years and above | **54%** | Strong concerns about unintended effects of the vaccine |
| El-Sokkary<br>2021 | Healthcare workers | **41,9%** | had not heard about Emergency Use Authorization (EUA) |

**Table 3.** Refusal to be vaccinated (rate and reasons)

| **Author** | **Participants** | **Rate of refusal** | **Reasons of refusal** |
| --- | --- | --- | --- |
| Adebisi , 2021 | Males and females who use social media over the age of 18 years old | **25.5%** | Unreliability of the clinical trials; belief that their immune system is sufficient to combat the virus |
| Ditekema 2021 | Adults 18 years of age and above |  | Did not trust the vaccine; believed the vaccine is made to kill people in Africa; believed the vaccine is made to sterilize people |
| Fares, 2021 | Healthcare workers | **28%** | The unknown protection and immunity duration and the rumours about the vaccine's available version; heard of anyone with a bad reaction related to COVID-19 vaccination; not trust pharmaceutical companies to produce a safe and effective vaccine and not believe that the side effects are discussed openly; |
| Saied, 2021 | Medical colleges students | **6%** | concerns regarding the vaccine's adverse effects and ineffectiveness; deficient data regarding the vaccine's adverse effects, and insufficient information regarding the vaccine itself |
| Acheampong 2021 | Adults | **21%** | uncertainty about the quality of the vaccine; insufficient information on the possible effects of the vaccine; and uncertainty about the vaccine's effectiveness in preventing them from contracting COVID-19 |
| El Kefi, 2021 | Staff of the Military<br>General | **31%** | Fear of side effects, doubts about the vaccine's efficacy, reluctance to any vaccination. |
| Oyekale, 2021 | General population | **6.6%** | Vaccine safety issues |
| El-Sokkary<br>2021 | Healthcare workers | **32.1%** | Had not heard about Emergency Use Authorization (EUA); perception for the severity of COVID-19 and COVID-19 vaccine safety |

### Risk of bias in studies

In terms of methodological quality, twenty studies were of good quality, two of moderate quality and one of poor quality.

### Results of individual studies

The articles included are from the following authors: Abebe et al [26] ; Acheampong et al [28] ; Adebisi et al [17]; Adeniyi et al [30] ; Aemro et al [31] ; Agyekum et al [32]; Angelo et al [34] ; Amuzie et al [33] ; Bongomin et al [18] ; Dinga et al [27] ; Ditekema et al [19] ; Echoru et al [20] ; El Kefi et al [35] ; El Sokkary et al [39] ; Elhadi et al [21] ; Fares et al [22] ; Handebo et al [23]; Kanyike et al [29]; Mose et al [24] ; Oduwole et al ; Omar et al [37] ; Oyekale et al [38] ; Saied et al [25]. Individual characteristics of the included studies are presented in tables 1, 2, and 3.

### Results of synthesis

#### Attitudes and reasons on COVID-19 vaccination in Africa

The results of the included studies indicate that the attitudes of people in Africa towards COVID-19 vaccines are diverse. These attitudes consist of hesitation, refusal and acceptance for various reasons.

#### Acceptance of vaccines

A total of nine studies reports vaccine acceptance rates varying from 6 to around 92%. ^[17–25,39]^ This acceptance of vaccination emanates from the general population aged 18 and over ^[26,28,36,37,19,17,21]^ and the population aged 18 to 70,^[20]^ medical students,^[25]^ health workers, ^[19,25, 27–32,35]^ women pregnant attending an antenatal clinic,^[25]^ primary and secondary school teachers ^[23]^ and patients attending outpatient clinics.^[18]^ Two studies report that people in the 18–20-year age group ^[20]^ and those over 46 ^[29]^ were inclined to accept the vaccine. In addition, male subjects were twice as likely to accept the vaccine; those who completed tertiary education, and students, Muslims, and the self-employed were more likely to accept the vaccine. ^[20]^ In addition, those attending secondary school and above, those with chronic illness and good knowledge of vaccines, were inclined to accept the vaccine. ^[26]^

Accepting vaccination is numerous and often specific to the type of study participants. The main ones are:

– the confidence in the accuracy of measures taken by the government in the fight against COVID-19,
– the fact that all vaccines available for COVID-19 are under the umbrella of Emergency Use Authorization (EUA), ^[39]^
– the fact that relatives have been diagnosed or have died with COVID-19, ^[31,18]^
– beliefs in the existence of COVID-19 ^[19]^ and being infected with COVID-19 at the time of vaccination, ^[21]^
– get vaccinated to protect family, friends and others in the community.^[28]^

In addition, the risks of being contaminated with COVID-19, the vaccine’s safety, the effectiveness of the vaccine, the facilitation of travel, the compatibility of vaccines with religion and being vaccinated would help strengthen immunity are also reasons that have motivated some people to accept the vaccination.^[22,29,32,34,35]^ Furthermore, some participants believe they have a public health responsibility to help fight the pandemic, and others felt that it is the community’s responsibility to get vaccinated.^[22]^ Speaking of responsibility, pregnant women must observe good practices about COVID-19 and its preventive measures, including vaccination.^[24]^ Moreover, it appears that the main factor likely to increase acceptance of vaccination is obtaining sufficient and precise information on available vaccines.^[22]^ Rates and reasons for vaccines acceptance are presented in table 1.

#### COVID-19 vaccine hesitancy

The rates of hesitancy to be vaccinated against COVID-19 reported by studies vary from around 1,06 to 85%. ^[17,28,20,22,25, 30–32,35–38]^ Study participants who say they are reluctant to get vaccinated against COVID-19 consist of the general population aged 18 and over, ^[17,27,28,35,36]^ adults aged 18 to 70, ^[20]^ healthcare workers ^[22,29–34,37]^ and medical school students.^[25]^ In addition, people aged 61 to 70, unemployed and pagan participants, the singles group and city dwellers were reluctant to accept the vaccine. ^[20]^

Study participants cite several reasons to justify their hesitation. For example, there is the fear of the quality of the vaccine distributed or sent to Africa, ^[27]^ the lack of sufficient clinical trials, insufficient information on the possible effects of the vaccine, uncertainty about the quality of the vaccine, uncertainty about the effectiveness of the vaccine in preventing them from contracting COVID-19,^[28]^ the unclear information provided by public health authorities, the low risk of contracting COVID-19 Infection, uncertainty regarding the tolerability of side effects of the vaccine,^[30]^ strong concerns about unintended effects of the vaccine ^[35]^ and the fear of side effects of the vaccine.^[22]^ Rates and reasons for hesitancy to get vaccinated are presented in the table 2.

#### Refusal to be vaccinated

The studies included in this review report refusal rates ranging from 6 to approximately 61%. ^[17,19,19,25,28,33,39]^ Medical students,^[25]^ healthcare workers ^[22,28,32,33,39]^ and adults aged 18 and over ^[17,19,28,35,36]^ were the participants in the studies that reported these refusals. Male participants, singles, and those who perceive a very high or moderate risk of contracting COVID-19 in the future are the most represented among those who refuse to be vaccinated. ^[29]^

The main reasons cited by those who refused COVID-19 vaccines are multiple. These reasons include the unreliability of clinical trials, the belief that their immune system is sufficient to fight the virus, ^[14]^ the lack of confidence in the vaccine and the lack of confidence in pharmaceutical companies to produce a safe and effective vaccine. ^[17,22,25,34,38]^ In addition, many claimed to have learned that all available vaccines for COVID-19 are under the Emergency Use Authorization (EUA) umbrella.^[34]^ Furthermore, concerns about the vaccine’s adverse effects and ineffectiveness have led some people to refuse vaccination. ^[22,28]^ In addition to these reasons, there are perceptions that the vaccine was intended to kill people in Africa, sterilize people ^[19]^ and doubt that the side effects are openly discussed. ^[22]^

Furthermore, the duration of protection and immunity is unknown, and rumors about the vaccine version are available. ^[22]^ Globally, the main obstacles to vaccination against COVID-19 are insufficient data regarding the vaccine’s adverse effects and insufficient information about the vaccine itself. ^[25]^ Rates and reasons for refusal to be vaccinated are presented in table 3.

## Discussion

This systematic review aimed to report attitudes and reasons associated with COVID-19 vaccination in Africa regardless of the study participants and design.

Three attitudes have been identified: vaccination acceptance, hesitancy, and refusal to be vaccinated. Several reasons have been associated with each attitude. Addressing these attitudes can help improve immunization coverage in Africa. Previous reviews have focused more on acceptance rates (intention), hesitation and refusal. However, they have not studied attitudes in detail to propose effective strategies.

Vaccination acceptance motivators were confidence in the government’s accuracy of measures to fight COVID-19 and that relatives have been diagnosed or died of COVID-19.

The hesitancy was justified by reasons such as fear of the quality of the vaccine being distributed or sent to Africa, the lack of sufficient clinical trials and fear of the vaccine’s side effects.

The refusal to be vaccinated was justified by reasons such as the unreliability of clinical trials, the belief that their immune system is sufficient to fight the virus, insufficient data regarding the vaccine’s adverse effects, and insufficient information regarding the vaccine itself.

The systematic review found that vaccine acceptance rates or willingness to be vaccinated against COVID-19 ranged from 6 to 92%. ^[17–25,39]^ Studies of Ebola vaccination have reported results of varying acceptance rates. Huo et al., ^[40]^ in their study titled “Knowledge and attitudes about Ebola Vaccine among the General Population in Sierra Leone”, reported that 72.5% of participants were willing to be vaccinated against Ebola if it was free. In a national household survey in Guinea, Irwin et al. ^[41]^ reported that 84.2% of participants said their family would accept Ebola vaccines. Always in Guinea, Kpanake et al. ^[42]^ found that 38% of participants said they were always ready to be vaccinated. Differences could be explained by study participants and the survey timing (before or during vaccination; before, during or after the epidemic).

In this systematic review, the reasons to get vaccinated against COVID-19 are similar to those for vaccination against Ebola. These reasons are mainly: the capacity of vaccination to prevent disease, the severity of the disease (its high case fatality rate), the safety of the vaccine, its efficacy and its availability. ^[27,21,22,30,31]^ Therefore, to convince people to get vaccinated against covid 19, the communication must be focused on the consequences of the disease, vaccine availability, its safety and effectiveness.

The rates of reluctance to be vaccinated against COVID-19 reported by studies vary from around 1,6 to 85%. ^[17,28,20,22,25,30–32,36–38]^ In their study on why the Guinean people were vaccinated against Ebola. Kpanake et al. ^[42]^ found a reluctance rate of 19%. This rate is a little low compared to the rates revealed by this systematic review. This could be explained by the nature of the pathologies and the effectiveness of communication actions.

The reasons for reluctance to be vaccinated are often fearful of the quality of the vaccine distributed or sent to Africa, ^[28]^ lack of sufficient clinical trials and fear of vaccine side effects. ^[22]^ The rapidity of vaccine circulation, poor communication, the withdrawal of certain vaccines, the abundance of debates and information could support the hesitation. If actions are not taken to reassure populations about the quality and side effects of vaccines, a large part of hesitant people could fall into the camp of those who refuse to be vaccinated.

The results of Huo et al. ^[40]^ Ebola vaccination study showed that 42% of participants did not intend to be vaccinated. In their study on the positions of the Guinean people to be vaccinated against Ebola, Kpanake et al. ^[32]^ reported that 25% of participants said they never wanted to be vaccinated. The results of these two studies range from 6 to about 61% ^[17,19,22,25,28,34,38]^ revealed by this systematic review. This difference could be explained by the diversity of research methods and the experimental or licensed nature of the vaccines.

Concerns about vaccine side effects, vaccine safety and efficacy ^[19,22,25,30]^ and the belief of study participants that their immune system is sufficient to fight the virus ^[20]^ are the most common reasons for refusing vaccinations. Therefore, it is essential to communicate about disease and vaccine effectiveness to avoid these false beliefs. It is also vital to build and strengthen trust with populations to ensure vaccination acceptance. ^[28,31,32]^ Unfortunately, this situation could lead to an increase in the rate of refusal to be vaccinated the persistence of the pandemic with a high case fatality rate.

### Strengths and limitations

The main strengths of this systematic review lie in the fact that it included all studies regardless of the design. In addition, the included studies looked at both the general population and specific groups. In addition, document searches were carried out in several databases and in gray literature to reduce the risks associated with publication bias.

There may be selection bias due to the restriction of publication languages. For example, the research looked at articles published in French or English. In addition, the synthesis was not conducted to have visibility of attitudes and reasons by type of participant. Furthermore, this review did not address the association of reasons with sociodemographic variables.

## Conclusion

Debates have been intense around COVID-19 vaccines since the start of vaccination campaigns. As a result, several studies have been carried out to determine people’s attitudes towards these vaccines. This systematic review looked at the attitudes of people in Africa towards COVID-19 vaccines. The included studies had the general population aged 18 and over and specific groups as participants. Three attitudes were reported at different rates; acceptance of vaccination, reluctance and refusal to be vaccinated. The safety, quality, side effects of vaccines and knowledge about COVID-19 are the main factors determining these attitudes. Therefore, this systematic review helps to understand the attitudes of the populations of Africa towards vaccines against COVID-19.

Meanwhile, the results indicate that research needs to focus more on identifying the facilitators of COVID-19 vaccination. This is to increase the acceptance rate significantly and, at the same time, reduce the hesitation and refusal rates. This research also provides essential elements for health personnel in charge of vaccination to develop strategies likely to obtain satisfactory coverage rates.

## Data Availability

All data produced in the present work are contained in the manuscript

